# Intra-host evolution during SARS-CoV-2 persistent infection

**DOI:** 10.1101/2020.11.13.20231217

**Authors:** Carolina M Voloch, Ronaldo da Silva F, Luiz G P de Almeida, Otavio J. Brustolini, Cynthia C Cardoso, Alexandra L Gerber, Ana Paula de C Guimarães, Isabela de Carvalho Leitão, Diana Mariani, Victor Akira Ota, Covid19-UFRJ Workgroup, LNCC-Workgroup, Cristiano X Lima, Mauro M Teixeira, Ana Carolina F Dias, Rafael Mello Galliez, Débora Souza Faffe, Luís Cristóvão Pôrto, Renato S Aguiar, Terezinha M P P Castiñeira, Orlando C. Ferreira, Amilcar Tanuri, Ana Tereza R de Vasconcelos

**Affiliations:** Departamento de Genética, Instituto de Biologia, Universidade Federal do Rio de Janeiro, Rio de Janeiro, Brazil; Laboratório de Bioinformática, Laboratório Nacional de Computação Científica, Petrópolis, Brazil; Instituto de Biofísica, Universidade Federal do Rio de Janeiro, Rio de Janeiro, Brazil; Departamento de Doenças Infecciosas e Parasitárias, Faculdade de Medicina, Universidade Federal do Rio de Janeiro, Rio de Janeiro, Brazil; Departamento de Cirurgia, Faculdade de Medicina, Universidade Federal de Minas Gerais, Belo Horizonte, Brazil; Simile Instituto de Imunologia Aplicada Ltda; Departamento de Bioquimica e Imunologia, Universidade Federal de Minas Gerais, Belo Horizonte, Brazil; Instituto de Biologia Roberto Alcântara Gomes, Universidade do Estado do Rio de Janeiro, Rio de Janeiro, Brazil; Departamento de Genética, Ecologia e Evolução, Instituto de Ciências Biológicas, Universidade Federal de Minas Gerais, Belo Horizonte, Brazil

**Keywords:** COVID-19, RNA-editing enzymes, quasispecies, Spike gene, Helicase gene

## Abstract

Prolonged infection of SARS-CoV-2 represents a challenge to the development of effective public health policies to control the COVID-19 pandemic. The reason why some people have persistent infection and how the virus survives for so long are still not fully understood. For this reason, we aimed to investigate the intra-host evolution of SARS-CoV-2 during persistent infection. Thirty-three patients who remained RT-PCR positive in the nasopharynx for at least 16 days were included in this study. Complete SARS-CoV-2 sequences were obtained for each patient at two time points. Phylogenetic, populational, and computational analysis of viral sequences confirmed persistent infection with evidence for a transmission cluster in health care professionals that shared the same workplace. A high number of missense variants targeting crucial structural and non-structural proteins such as Spike and Helicase was found. Interestingly, longitudinal acquisition of substitutions in Spike protein mapped many SARS-CoV-2 predicted T cell epitopes. Furthermore, the mutational profiles observed were suggestive of RNA editing enzyme activities, indicating innate immune mechanisms of the host cell. Viral quasispecies analysis corroborates persistent infection mainly by increasing richness and nucleotide diversity over time. Altogether, our findings highlight a dynamic and complex landscape of host and pathogen interaction during persistent infection suggesting that the host’s innate immunity shapes the increase of intra-host diversity with possible implications for therapeutic strategies and public health decisions during the COVID-19 pandemic.

## Introduction

The disease caused by the new coronavirus SARS-CoV-2 (COVID-19) was first reported in December of 2019 and has rapidly spread worldwide, in a pandemic that already surpasses 51,805,339 cases and 1,279,184 deaths. In Brazil, the Coronavirus Resource Center reported more than 5,7 million cases and 162,842 deaths between March and November (https://coronavirus.jhu.edu/map.html), and active transmission is still observed all over the country.

Observational studies conducted since the beginning of the pandemic have been crucial to providing a detailed description of the main demographic and clinical characteristics associated with SARS-CoV-2 infection in different populations (Grasselli et al. 2020; Richardson et al. 2020; Zhou et al. 2020). Most of these studies were hospital-based and, consequently, the natural history of SARS-CoV-2 infection among mild cases has been less documented. However, a description of mild cases can be considered crucial to determine transmission patterns and better define the minimum time of in-home isolation and other preventive policies.

The medium time between symptoms onset and the first negative RT-PCR test has been described as 15 to 17 days in nasopharyngeal swabs, although longer periods are often observed, varying according to the clinical specimen under analysis (Pavon et al. 2020; Sun et al. 2020; Xu et al. 2020). Also, cases of reinfection by phylogenetically distinct SARS-CoV-2 viruses were recently reported (Prado-Vivar et al. 2020; To et al. 2020). These data raise the question of whether the virus may establish persistent infection or if patients presenting prolonged viremia in the nasopharynx have been re-infected before recovering from the initial infection.

Among hospitalized patients, the criteria for discharge has been defined as resolution of symptoms in addition to two consecutive negative SARS-CoV-2 RT-PCR tests collected in a 24h interval (CDC 2020a; ECDC 2020). By contrast, symptom-based criteria for discontinuation of in-home isolation have been defined, including improvement in clinical symptoms and at least 24 hours since last fever without the use of antipyretic medication (CDC 2020b). Hence, patients with mild infection are usually released from in-home isolation after resolving clinical symptoms without proper testing to confirm viral clearance. Returning to work without testing can be particularly dangerous to the susceptible population in contact with health care workers due to their high exposure in the hospital environment.

In brief, persistent infection with SARS-CoV-2 has been characterized mainly by prolonged PCR positivity. However, studies available to date did not include viral sequencing or a detailed analysis of viral infectiousness or intra-host evolution, which would be expected in such cases. Indeed, we still don’t know the clinical outcome associated with persistent infection. Here, we investigated a series of 33 patients from the cities of Rio de Janeiro and Belo Horizonte (Brazil), who remained RT-PCR positive for at least 16 days after the first positive test. Samples were obtained from each patient at two time points. Phylogenetic, populational, and computational analysis of SARS-CoV-2 sequences confirmed persistent infections and showed increasing diversity associated with persistence, presenting immune escape related mutations and editing signatures of APOBEC induced interferon enzyme.

## Results

### Study cohort

Twenty-one females and 12 males with mean ages of 39 ± 11 and 38 ± 9 years, respectively, were enrolled in this study. Most participants were health professionals (N = 19) or other workers (N = 7) from hospitals and clinics from the city of Rio de Janeiro. A single patient was recruited from the State of Minas Gerais (Table S1). Patients from Rio de Janeiro were selected from a larger cohort of 4,917 individuals, with an estimated prevalence of 30% of persistent SARS-CoV-2 infection. We defined viral persistency in our cohort as patients that remained positive for SARS-CoV-2 in nasopharyngeal samples for at least 14 days since the onset of the symptoms. We sequenced samples collected at two time points for each patient (T1 and T2). T1 was the first RT-PCR positive sample after the onset of symptoms, and T2 the last sample with a Ct < 35, which is required for sequencing. The mean interval between the two samples was 18±7 days (range 5 - 39). The longest interval between T1 and the onset of the symptoms was 22 days (**Table S1; Fig. 1A**).

**Figure 1.**
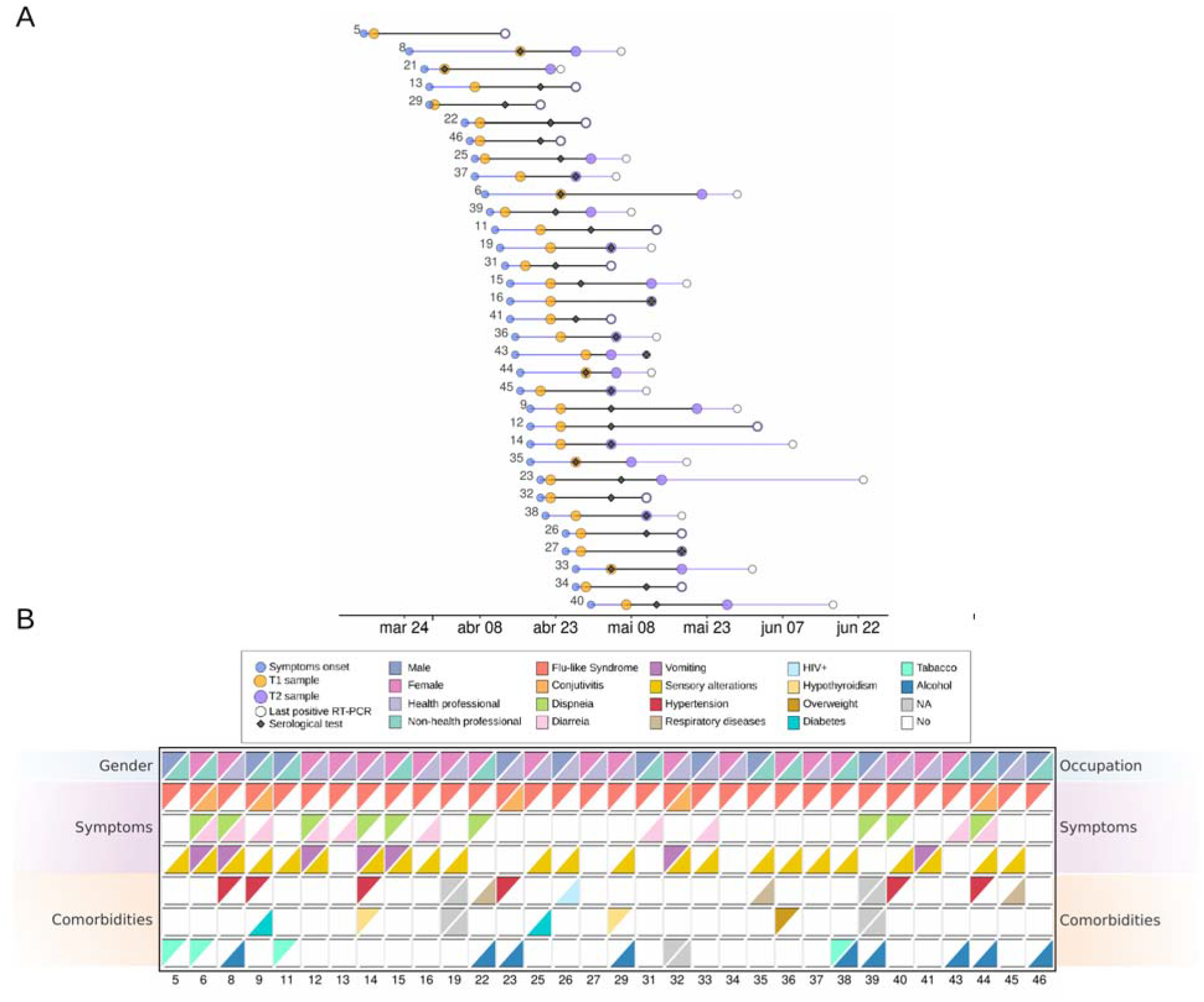
Characterization of patients with persistent SARS-CoV-2 infection. **A)** Time intervals between onset of patients’ symptoms, first sample sequenced, first positive serological test, second sample sequenced, and last positive RT-PCR test, respectively. Blue lines represent the interval between onset of patients’ symptoms and first sample sequenced. Black lines indicate the difference between the two samples sequenced whereas in purple we show the difference between the second time sample and the last positive RT-PCR test. **B)** Key clinical features for the patients analyzed in this study. Each row indicates two different features, first row: gender and occupation; second-to-fourth rows: symptoms, fifth-to-seventh rows: comorbidities. Patients are represented in the columns.

Fifteen patients did not declare any pre-existing health condition, and hypertension was the most prevalent comorbidity (N = 6) among those who declared. All patients developed mild, flu-like symptoms of COVID-19. Sensory changes, diarrhea, and vomiting were also observed in a few participants (**Fig. 1B**). A single patient, who reported hypertension and hypothyroidism, required hospitalization for two days. As expected, the Ct values were higher in T2 when compared to T1 (**Table S1**). Serology tests were also performed, and all patients were reagent for ELISA anti-SARS-CoV-2 spike protein at T2. Six patients were already reagent at T1 (**Fig. 1A**).

### Phylogenetic analysis of consensus genomes

Reference-based viral genome assembly achieved, on average, 97% genome coverage with a read depth greater than 2,000x in most of the regions sequenced (**Table S2**). Similar results were found using a *de novo* assembly approach. To analyze the phylogenetic position of the 66 consensus genomes generated in this study, we put together a dataset including 135 SARS-CoV-2 Brazilian genomes from Rio de Janeiro, São Paulo, and Minas Gerais states obtained from samples collected between March and June of 2020. The maximum likelihood tree estimated for these 201 genomes is shown in **Fig. 2**. The analyzed sequences were classified by Pangolin software into seven distinct lineages recovered in well-supported clades. The majority of our genomes (58 out of 66) were classified as belonging to lineage B.1.1.33. Only two were classified as B.1.1.28, four as B.2.2, and two as B.1. For all patients, T1 and T2 genomes were always classified as belonging to the same lineage (**Fig. 2**).

**Figure 2.**
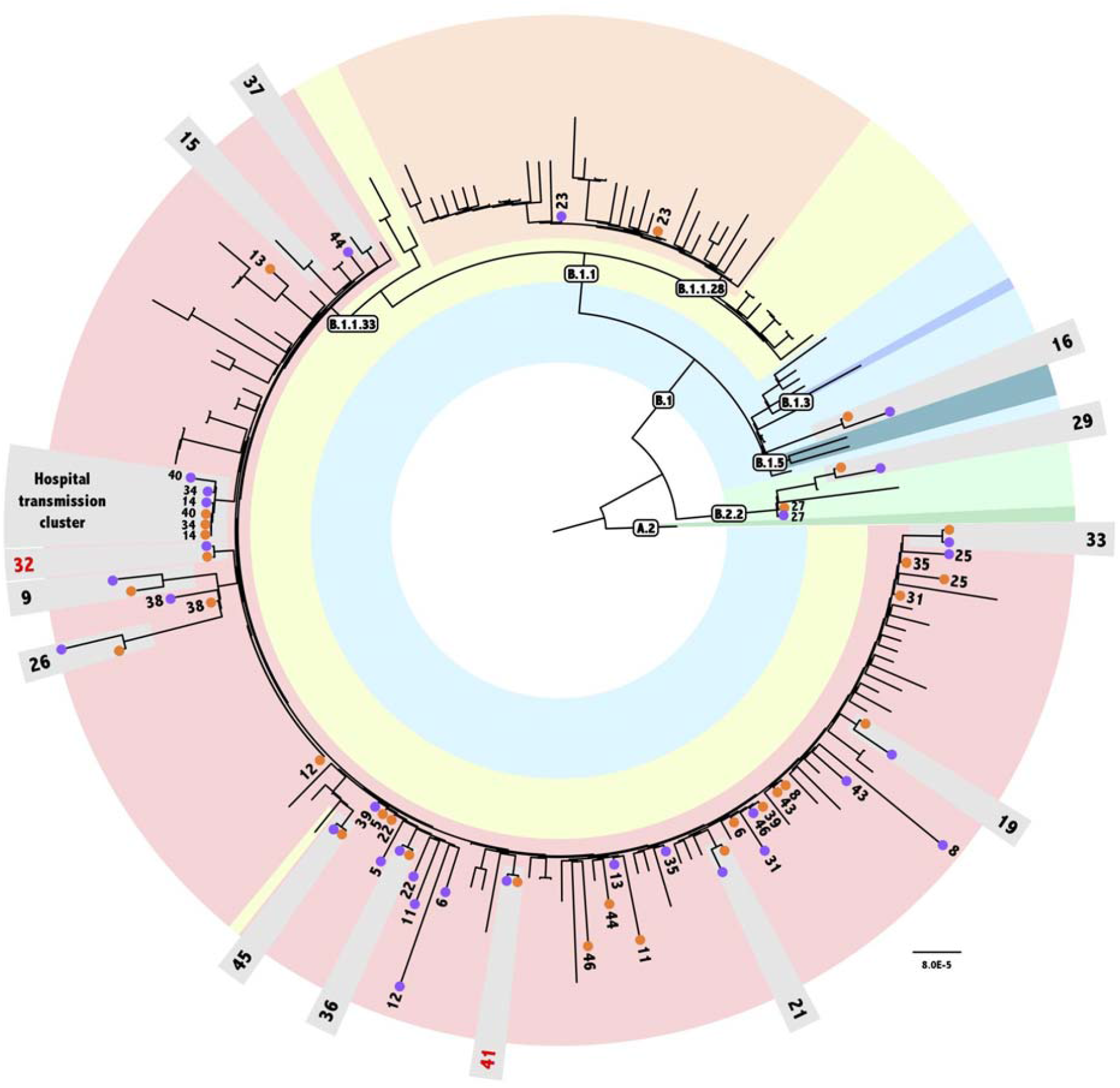
Phylogenetic analysis of persistent samples. Maximum Likelihood tree obtained with Consensus dataset analysis under GTR+I model, containing a consensus genome for each time point of the 33 patients plus 135 populational samples. Populational sample names were excluded from the figure for clarity. Text boxes indicate the Pangolin lineage classification. A different color represents each sampled lineage in the tree. Numbers indicate each patient’s samples. Orange and purple circles represent T1 and T2 samples, respectively. Patients whose the two time point samples clustered are highlighted in light-grey boxes. Red numbers indicate patients that are not monophyletic in the haplotypes tree.

Phylogenetic analysis revealed the monophyletic status of the pair of samples (T1 and T2) obtained from 13 out of the 33 patients analyzed in this study, namely, patients 9, 15, 16, 19, 21, 26, 29, 32, 33, 36, 37, 41 and 45 (**Fig. 2**). Ancestral sequence reconstruction indicates that the number of nucleotide substitutions in the branches supporting the monophyly of these patients vary between one and five. Taking into account the diversity of the GISAID genomes analyzed in this dataset, ancestral state reconstruction accurately inferred substitutions associated with the characterization of lineages defined by Pangolin, highlighting the robustness of our analysis. Therefore, we tried to understand why some genome pairs from the same patient did not cluster in our tree. Our estimated tree has 169 near-zero length internal branches, and all these pairs are separated only by those branches. Indeed, when comparing each near-zero branch with its immediate ancestor, ancestral state reconstruction did not attribute any well-supported substitution within the whole genome for any of these near-zero branches. Therefore, considering that all patients enrolled in the study remained in social isolation until they had a negative PCR result and that their consensus sequences are more equally related to each other than to any other virus in the population, we have no evidence to consider the reinfection of those patients.

We also found evidence of a transmission cluster for patients 14, 34, and 40, who are health professionals at the same hospital (**Fig. 2**). All genomes (T1 and T2) from those patients share two substitutions, U175C and G2086U (ORF1ab). Patient 14 presented symptoms first followed by 34 and 40, with a T1 sampling interval of 5 and 13 days, respectively. Indeed, T1 consensus sequences from these three patients, as well as T2 sequences from patients 14 and 34 were identical. Patient 40’s T2 sample harbored only one substitution (U22119C) when compared to the other five consensus sequences previously described. Since the probability of having this shared variation as a result of recurrent mutations is too small, our results strongly suggest that they are the result of transmission between patients, indicating different events of infection with the same circulating virus in the hospital environment.

### Intra-host viral diversity and genetic characterization

Genetic screening across the 66 genomes revealed 327 intra-host single-nucleotide variants (iSNVs) in 304 positions spread throughout 9 ORFs of the SARS-CoV-2 genome (**Table S3**). Non-synonymous changes comprehend 221 out of 312 coding region variations (**Fig. 3A**). Orf1ab harbored the majority of the iSNVs detected in our analysis (N = 203) followed by the S protein (46), N (31), orf3a (12), M (5), orf7a (5), orf8 (5), orf6 (3), E (2), and non-coding regions (15). Nevertheless, when normalized by gene length, orfs 3a, 7a, 8, and 6 showed the highest proportion of variation.

**Figure 3.**
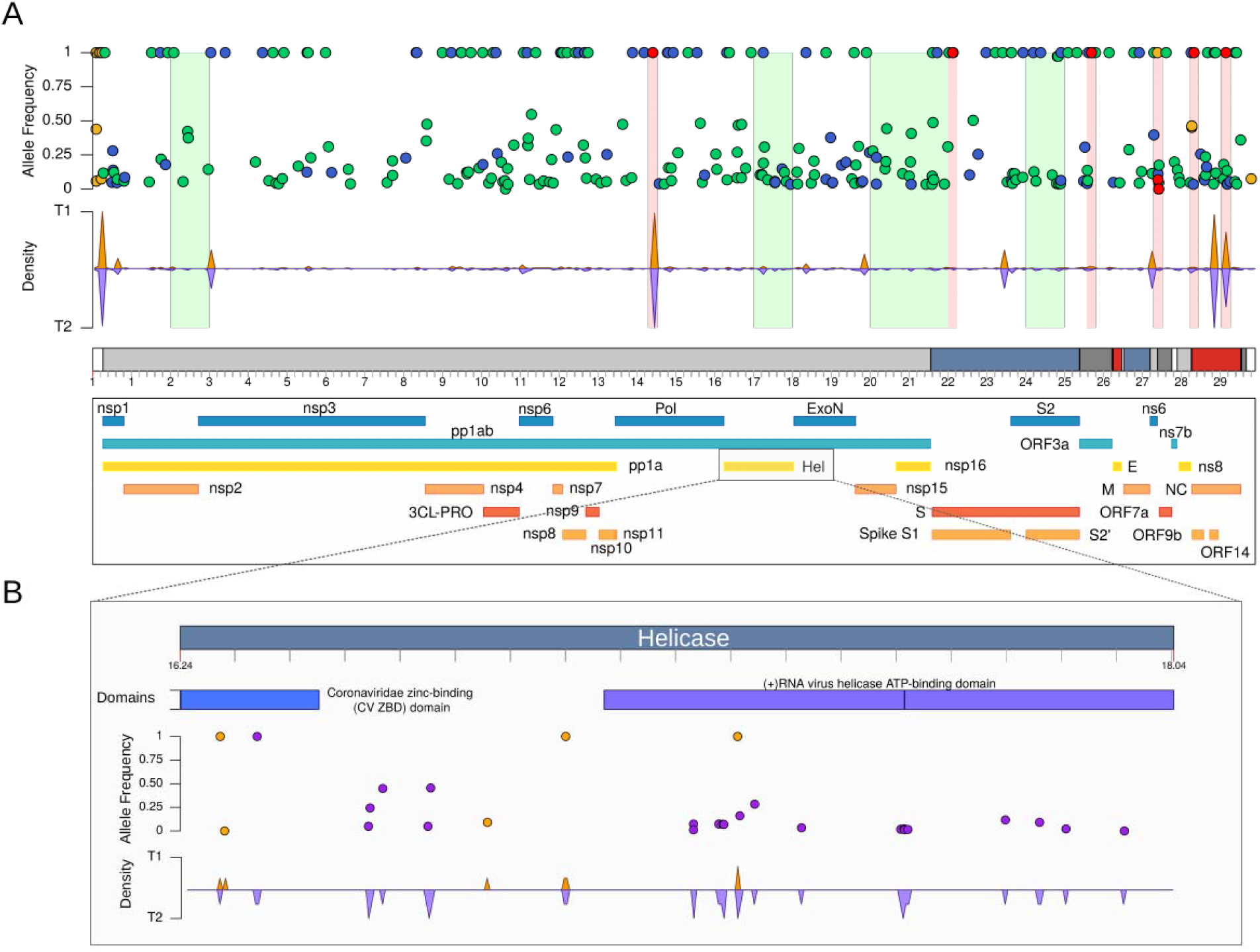
Genomic variation across SARS-CoV-2 genomes. **A) (Top)** intra-host Single Nucleotide Variant (iSNV) dispersion. Blue and green points indicate synonymous and non-synonymous mutations, respectively. In yellow, we show variants in non-coding regions. Red points represent non-synonymous sites under positive selection according to MEME. **(Middle)** Density plot showing the accumulation of iSNVs across virus genome in orange (T1 - first sample sequenced) and in purple (T2 - second sample sequenced). **(Bottom)** Protein product translated by SARS-CoV-2 ORFs. **(B)** Zoom in helicase (nsp13) protein showing differential accumulation of genetic variants in T2 samples when compared with T1. Variants were mostly spread across the ATP binding domain.

Intra-host estimates of non-synonymous substitutions may be excessively high because some sequence variation is attributed to mutations that have not been filtered by selection. To test the selection hypothesis, we performed a diversifying selection analysis using the MEME method implemented in Datamonkey. We detected seven individual sites subject to episodic diversifying selection considering a p-value threshold of 0.1 (genome position: 14407-14409, 22118-22120, 25690-25692, 27400-27402, 27418-27420, 28331-28333, 29147-29149) (**Fig. 3A**). These sites fall within ORF1ab, S, ORF3a, ORF7a, and N. Interestingly, only one of them, 14407-14409 (ORF1ab), had already been detected by the Datamonkey SARS-CoV-2 natural selection analysis server (available at http://covid19.datamonkey.org/), and is the one that was inferred to have more branches under selection in our analysis. Furthermore, we noticed a significant increase in the ratio of non-synonymous/synonymous substitutions (Wilcoxon test, *P* = 0.043; **Fig. S1A**) between T1 (mean = 2.22) and T2 (mean = 3.14) samples. Such differences are mainly due to the enrichment of missense variants at orf1ab (Fisher’s exact test, 2 × 2 contingency table, *P* = 0.0099).

Overall, we observed only one substitution per site across all variants called in the SARS-CoV-2 genome. However, we detected 21 two-state polymorphic sites, and less often, three alternative bases were identified per site (**Table S3**). Most of these sites mapped to orf1ab (12), S (4), orf3a (2), N (2), and one in the 5’ UTR. The co-occurrence of genetic variants was mostly found intra-host; however, nine sites were over-represented in more than 80% of samples. These positions correspond to mutational signatures of the most prevalent circulating lineage, namely B.1.1.33.

By comparing the mean number of iSNVs between the first (mean = 14.8) and second (mean = 16.4) time points during the course of infection, we observed a significantly higher number of iSNVs at the T2 sampling (Wilcoxon test, *P* = 0.005; **Fig. S1A**). Of note, we observed an elevated number of iSNV at T2 of patient 31 (n = 53), which exceeded more than 3 fold the expected mean. On average, each patient shared 10 variants between both time points, approximately three variants were only found in T1 and seven gained in T2. Overall, the T2 sample exhibited more exclusive variants than T1. A total of 81 variant sites were found only in T1, whereas 218 were acquired later in T2 (**Table S4**).

To rule out NGS data sequencing and processing steps as a spurious source of variation, we tested the correlation between our findings and quality control metrics. The difference between iSNV numbers across both times was found to be unrelated to genomic coverage (Spearman’s correlation ρ=-0.042, *P* = 0.7382). However, the number of iSNVs was positively correlated with the sampling interval from symptoms onset (Spearman’s correlation ρ = 0.41, *P* = 0.0006) and inversely proportional to RT-PCR Ct values (Spearman’s correlation ρ = −0.36, *P* = 0.0031). Altogether, this suggests that increasing variation at T2 could be explained by mutation accumulation during a more extended period of infection and persistence.

We then estimated the magnitude of difference between T1 and T2 samples by comparing the rates of iSNVs per kilobase (kb) and per protein product along the SARS-CoV-2 genome. Both approaches revealed an elevated accumulation of variants (LogFC > 2) overtime at seven main genomic windows, predominantly enriched at the 3’ UTR (**Table S5**). Interestingly, these regions are responsible for encoding Helicase, nsp15, nsp16, Spike subunit S2, nsp2-nsp3, M, and ns8 proteins, respectively. Alterations in Helicase protein mainly affected the ATP binding domain that donates the energy necessary to solve RNA secondary structures required during virus replication (**Fig. 3B**).

### Characterization of SARS-CoV-2 spike protein T cell reactive and predicted epitopes

To explore the impact of spike variations on the host immune response or escape mutants, we intersected the iSNVs detected in S with known and predicted T cell epitopes. We found 11 iSNVs intersecting ten known S-reactive CD4+ T cell peptides in eight different patients (**Fig. 4; Table S6**). Nine out of the 11 variants had a non-synonymous effect being mostly found in T2 samples, while only patient 37 harbored a variant in the T1 sample. Patients 16 and 25 had two and three variants mapping at least one reactive epitope, respectively. In addition, cross-referencing comparison with predicted CD4+ and CD8+ T cells revealed seven iSNVs overlapping six different T CD4+ epitopes (**Fig. 4; Table S6**). Alterations in T CD8+ predicted epitopes were reported in 28 peptides in 62 samples (**Table S6**). The overwhelming majority of patients harbored the D614G substitution mapped in the YQDVNCTEV epitope. This peptide was also predicted as a putative HLA class I candidate binding peptide, together with 31 other regions with a high probability for immune recognition of SARS-CoV-2. Finally, 28 HLA class II candidate peptides had at least one variation across 27 patients (**Table S6**).

**Figure 4.**
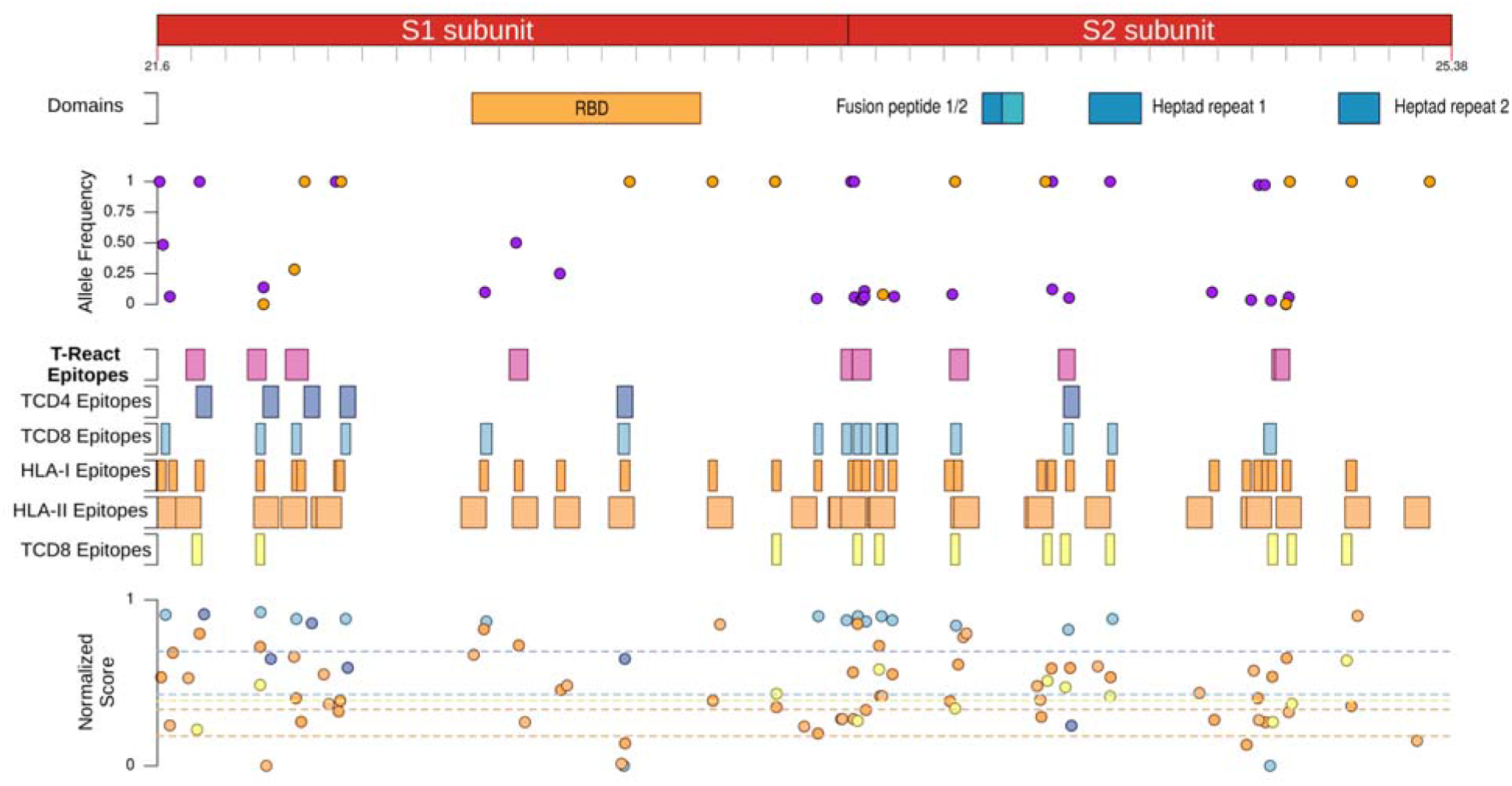
Spike protein variant characterization. Zoom in spike protein showing (**Top**) variants found in T1 samples in orange circles and variants in T2 in purple. (**Middle**) Density plot for each time and T cell (i) reactive epitopes, (ii) IEDB T CD4+ predicted epitopes, (ii) NetMHCpan4.0EL T CD8+ predicted epitopes, (iv) HLA-I candidate binding peptides, (v) HLA-II candidate binding peptides and (vi) T CD8+ epitopes predicted by Rosetta. Both regions harbored at least one iSNV in our patients. (**Bottom**) Normalized score for each predicted epitope represented by filled circles. Dashed lines represent the mean value of the score assumed by the S-reactive epitope in each computational approach considering the color matching.

### Dynamic alterations of mutational signatures in SARS-CoV-2 persistent infection

Next, we examined the occurrence of differential mutational profiles in the virus genome as a possible factor associated with persistent infection. Viral sequences exhibited a dynamic scenario of base substitution rates mainly dominated by C>U (42%), G>U (11%) and U>C (10%) variations. Although less common, longitudinal acquisition of A>U transversions characterized samples in the second time point of our analysis (Wilcoxon test, *P* = 0.006; **Fig. S1B**). On the other hand, the proportion of C>A and G>A significantly decreased over time (Wilcoxon test, P < 0.05; **Fig. S1B; Fig. S2**). Multiple pairwise correlation tests showed that the decrease in G>A is related to the G>C mutations and negatively related to A<->U signatures, which may indicate that both arose in a dependent manner (**Fig. S3**). In addition, U<->C mutations also showed an inversely proportional relationship (**Fig. S3**).

We built a classification model using relevant features that had been previously highlighted in our analysis and might distinguish long-term infection samples. ADA Boost classification demonstrated that T1 and T2 genomes could be distinguished with 79,84% accuracy. The ROC curve with AUC of 0.9, 84% sensitivity, and 87% specificity (**Fig. S4**) demonstrated a slight difference between the classified groups. This difference could be understood by analyzing the feature importance of the classification model (**Fig. S4**). The most important features in T1 and T2 are related to viral load, missense variants, total variants and G>A signature. The F1 score metric reached 86% which indicates a balanced relation between precision and recall. The 84% sensitivity suggests a slight difference in the classification of T1 and T2. Since the difference is minor when sensitivity is compared with specificity, we considered the model’s performance acceptable for classification. The variance structure accurately shows the most important feature of the classification model.

### The complexity of viral quasispecies composition mediates SARS-CoV-2 adaptability

To gain insights into viral population structure and dynamics during persistent infection, we performed a per-sample haplotype reconstruction using high and low-frequency variants. A total of 182 viral quasispecies were reconstructed with an average of 3 entities per sample. Pangolin algorithm classified all haplotypes as having the same lineage as the consensus lineages of their patients. Phylogenetic inference of the reconstructed haplotypes was mostly in agreement with the consensus tree (**Fig. 5**). Pangolin lineages were also recovered in well-supported clades. Patients whose T1 and T2 genomes clustered in the consensus analysis were also monophyletic in the haplotype phylogenetic reconstruction, with exception to patients 23, 32, and 41.

**Figure 5.**
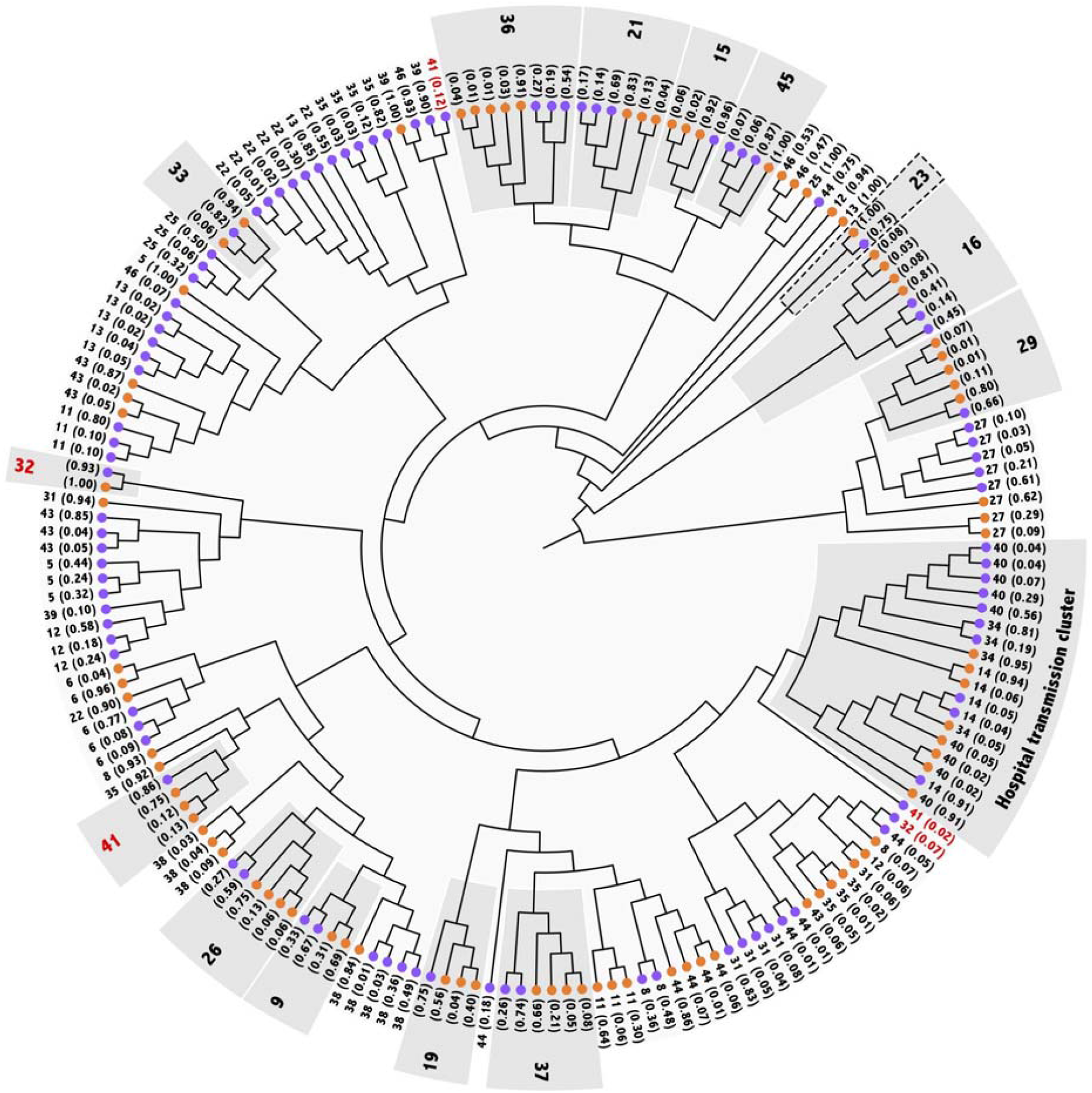
Haplotypes phylogenetic analysis. Unscaled ML (TN+R2) tree depicting phylogenetic relations of inferred haplotypes for the two time point samples of the 33 patients. Orange and purple circles represent T1 and T2 samples, respectively. Patients are identified by the number, and the estimated frequency of each haplotype is between parenthesis. Light-grey boxes highlight patients whose consensus sequences clustered in the consensus tree. Numbers in red indicate that low-frequency haplotypes are not monophyletic with the respective patient’s main clade.

Particularly, T1 and T2 genomes from patient 23 did not cluster in the consensus tree and were monophyletic in the haplotype phylogenetic inference. However, inferred haplotypes clustered due to the absence of other samples from the B.1.1.28 lineage. For patients 32 and 41, the most frequent haplotypes were monophyletic but at least one minority haplotype did not cluster. Similarly, the separation of these minority haplotypes from the monophyletic clade formed by other haplotypes from the same patient is not well supported by bootstrap values. This should be interpreted as a lack of difference between haplotypes rather than as a sign of distinction between them. Indeed, we inferred an identical haplotype as the most frequent for 13 patients. Namely, patients 5, 6, 8, 12, 13, 22, 31, 35, 39, 41, 43, 44 and 46 presented the same haplotype for at least one sample. Patients 5, 6, 8, 12, 31 and 43 presented that inferred haplotype for T1 samples while patients 13, 41, 44 and 46 for T2 samples and, finally, patients 22, 35 and 39 presented an identical haplotype in samples T1 and T2. Low-frequency haplotypes were mostly found to be exclusive to each sample.

The same pattern was observed among patients 14, 34 and 40 that included the hospital staff cluster. The most frequent haplotype is the same in all T1 samples, as well as in T2 samples from patients 14 and 34, corroborating the transmission hypothesis that we have proposed from the consensus sequence analysis. Interestingly, a low frequency iSNV (C622A) is present in T2 sample from patient 14, and in T1 from patients 34 and 40. This suggests that Patient 14, who was the first of the three to develop symptoms, contaminated patients 34 and 40, and that the transmitted virus included some low frequency iSNVs, along with the most frequent ones.

No statistically significant difference was found between the number of haplotypes at both time points (**Fig. S5**). Nevertheless, within-sample polymorphic sites (i.e., mutated positions compared to dominant haplotype) were enriched in T2 samples (Wilcoxon test, *P* = 0.003; **Fig. S5**). Abundance-based diversity analysis revealed an important increase in normalized Shannon entropy in T2 samples (Wilcoxon test, *P* = 0.029, **Fig. S5**). Quasispecies abundance in T1 was characterized by the presence of one dominant haplotype with a frequency greater than 80% in most of the cases, whereas in T2 this frequency decreased to nearly 50% (**Fig. S6**). Although the number of haplotypes is comparable, discordant nucleotide sites within quasispecies were smaller in T1 samples (**Fig. S5**). Consequently, these samples were more related considering both the sum of pairwise distances between haplotypes and nucleotide diversity. Therefore, amplitude of the SARS-CoV-2 mutant spectrum during persistent infection is mainly recognized by differential richness and extensive distribution of variants.

## Discussion

Prolonged SARS-CoV-2 infection represents a great challenge to the development of effective public health policies to control the COVID-19 pandemic. Due to the low availability of PCR tests, there is a prevalent use of symptom-based criteria for interruption of primary care management and self-isolation, usually two weeks since the onset of symptoms. Prevalence of persistent COVID-19 symptoms affect between 10-30% of patients worldwide and may last 3-12 weeks (Greenhalgh et al. 2020; Ladds et al.). The high prevalence of persistent cases indicates that this is not a rare phenomenon and needs to be explored to better control epidemic spread. Nevertheless, the reason why some people have persistent infection and how the virus survives for so long are still not fully understood. Indeed, persistent infection has been described for many other respiratory viruses, including influenza (Wang et al. 2018), MERS (Arabi et al. 2018), and RSV (Gomez 2012). In this study, we successfully identified patients with persistent SARS-CoV-2 infections by performing a longitudinal study of viral intra-host genomic evolution using a high-throughput sequencing approach in 33 patients infected for over two weeks. Our results demonstrate that SARS-CoV-2 may establish persistent infection in the nasopharynx, even in seroconverted patients, indicating that the safest way to reduce the transmission of COVID-19 is through PCR testing to ensure viral clearance.

The phylogenetic analysis allowed the undeniable identification of 13 patients with long-term infection due to the monophyletic grouping between T1 and T2 consensus sequences. However, even with growing evidence that SARS-CoV-2 reinfections may occur (Parry 2020; To et al. 2020), we have no clear indication that supports the hypothesis of reinfection in our cohort. We detected patients with persistent infection from four different lineages suggesting that long-term infections are not exclusive from a unique lineage. None of our patients had an inconsistent lineage assignment in T1 and T2, reinforcing the exclusion of re-infection events in our cohort. Candido et al. (2020) showed that community transmission of COVID-19 in Brazil was established from limited clades. Therefore, until now, little variability is observed between most of the virus circulating in the country. This situation justifies the expressive number of near-zero branches and the lack of well-supported substitutions in the ancestral state reconstruction, close to the base of each lineage in the tree. In the absence of a well-supported branch or a state change in the inferred ancestry, we cannot confirm any case of reinfection.

Returning to work without testing can be particularly dangerous among healthcare workers due to their high exposure and contact with the susceptible population. In the analyzed cohort, mainly composed of healthcare workers, long-term infections represent 30% of cases. This extremely high frequency of persistent infections may be a caveat of our cohort. Populational studies including asymptomatic individuals in a randomized analysis could lower this proportion. Of note, we have amplified and sequenced the full-length genome of viruses in T2 samples from these patients. The full-length genomes sequenced had all SARS-CoV-2 ORFs intact and consequently, are possibly able to infect cells and transmit to other persons.

In fact, the transmission cluster formed by health care workers of the same hospital highlights the preservation of the infectious ability of the virus in T2 samples. Moreover, there is a 13 days interval from the first patient’s symptom onset to that of the last patient, demonstrating the maintenance of the virus’s ability to establish successful prolonged infection events even after large intervals of time. The finding of transmission of intra-host variants between patients of the cluster, meaning that infection doses were large enough to contain low-frequency iSNVs, is essential to understand the nature of SARS-CoV-2 transmission and needs further investigation. Previous studies also reported the spread of the SARS-CoV-2 virus within hospital workers providing evidence for maintaining the prolonged infectious state (Rivett et al. 2020; Sikkema et al. 2020; Suárez-García et al. 2020).

From the viewpoint of mutational signatures, our findings may reflect host RNA-editing enzyme activities on the viral genome as a cell defense mechanism. For instance, high levels of C>U described here and reported by several authors may be caused due to APOBEC-mediated deamination, whereas G>U and U>C have been previously associated with RdRp error mutational spectrums (Smith et al. 2013). Of interest, G>A was also recently reported as part of the editing signature of the APOBEC-family (Niavarani et al. 2015). G>A was an important feature to classify persistent infection samples according to our machine learning model. ADA Boost also separates the two time points of infections using other variables such as viral load, missense variants and other mutational signatures with high sensitivity and specificity according to AUC of the ROC curve and F1 score. We noticed a decrease in G>A and C>A mutations in T2 samples possibly because of nonsense-mediated decay pathway recognition of premature stop codon induced by APOBEC editing (Chester et al. 2003). Adenosine sites are also target of A-to-I editing meditated by the ADAR enzyme. Thus, we cannot exclude the possibility of fewer adenosine mutated sites at the second time point mediated by ADAR activity, which seems to be more effective in restricting viral propagation than APOBEC (Di Giorgio et al. 2020). Both RNA editing enzymes have already been described as antiviral factors stimulated by interferon in many other RNA viruses.

Structural and non-structural proteins such as S and Helicase appear to play an important role in this process based on differential accumulation of variants. Helicase is a conserved protein responsible for the resolution of RNA secondary structures during the replication cycle of the virus (Jia et al. 2019). Thus, targeting helicase activity through the use of inhibitors is a potential candidate for COVID-19 therapy (Habtemariam et al. 2020). Mutations in the Helicase protein were previously correlated with D614G in S protein and impact virus infectivity (Kannan et al. 2020). Recent findings showed that D614G mutation increases the virus fitness and is associated with higher viral loads in the upper airway of COVID-19 in patients and infected hamsters suggesting the role of D614G mutation in viral transmissibility (Plante et al. 2020). In clinics, patients with G614 mutation developed higher levels of viral RNA in nasopharyngeal swabs than those with D614 virus but did not develop severe disease (Lorenzo-Redondo et al. 2020). Indeed, competition and functional analysis showed that viruses with the G614 signature overcame the original virus even in excess of D614. In fact, the G614 is characteristic of B.1 lineage and, consequently, is present in all genomes of this lineage and sub-lineages. Although most of our patients harbored the D614G mutation, which might contribute to its persistence in upper airway samples, such mutation is absent in patients 27 and 29, which belong to B.2.2 lineage. From all Brazilian genomes sequenced so far, more than 96% have this signature, a portion that is much higher than expected for persistent infections (Greenhalgh et al. 2020; Ladds et al.). Altogether, this indicates that more than the presence of D614G mutation, there is an alternative mechanism underlying the persistence in these patients.

Genomic variations are the main source of molecular evolution and adaptability in many RNA viruses. We demonstrated that SARS-CoV-2 intra-host genetic diversity is a possible cause of persistent infection. Quasispecies diversity provides the virus a greater chance to evolve and adapt during infection (Eigen 1993; Coffin 1995; Domingo et al. 1996). Intra-host diversity is implicated with pathogenicity and infection outcomes for many other viruses (Vignuzzi et al. 2006; McWilliam Leitch and McLauchlan 2013; Ni et al. 2016). According to the IEDB database, some of the mutated antigen sequences identified in our analysis may have differences in binding affinity for epitopes to MHC-I and MHC-II, once most of iSNVs in Spike protein mapped to T cell predicted epitopes. Therefore, these variations could be associated with a mechanism of escaping the host’s immune response and should be further investigated for the development of antiviral therapy and vaccines.

In conclusion, our study sheds light on the routes for intra-host genomic evolution of SARS-CoV-2 during persistent infection. We observed that most intra-host variations in SARS-CoV-2 present individual specificity and were not longitudinally transmitted, indicating that they probably were not adaptative. The RNA-editing enzyme activities of the innate immune system of the human host can be associated with the temporal accumulation of iSNVs along the SARS-CoV-2 genome, and this generated quasispecies diversity may be correlated with the outcome of persistent infection. Whether the upregulation in the mutation rate of Spike and Helicase is an adaptive feature still needs further investigation. Our findings have potentially exploitable implications for public health decisions during the management of the COVID-19 pandemic as well as therapeutic uses that should be investigated.

## Materials and Methods

### Study participants and sample collection

Thirty-three individuals with persistent infection by SARS-CoV-2 were enrolled in the study. Subjects from both genders were recruited at the Center for COVID-19 diagnosis from the Federal University of Rio de Janeiro and Simile Medicina Diagnóstica at Belo Horizonte from March to June, 2020. Persistent cases were defined as those who remained positive for SARS-CoV-2 in nasopharyngeal samples for at least 14 days since the onset of symptoms. Detection of SARS-CoV-2 and human RNase P were performed by RT-PCR using the CDC protocol (Waggoner et al.). Blood samples and nasopharyngeal swabs were obtained from each patient at two time points, and time 1 (T1) was determined as the first sample with a positive RT-PCR test. Only samples with a Ct < 35 were sequenced. Clinical and demographic data were self-reported by the patients. The present study was approved by the National Commission of Ethics in Research (protocol numbers 30161620.0.0000.5257 and 30127020.0.0000.0068). Written informed consent was obtained from all participants.

### Enzyme-linked immunosorbent assay

Microtitre plates (Immulon 2 HB) were coated with a trimeric spike proteins at 4 μg per ml in 100 mM sodium carbonate-bicarbonate buffer (pH 9.6) and incubated overnight at 4°C. Excess protein was removed by washing five times with PBS + 0,05% Tween-20 (PBST, Sigma) and unbound sites blocked with 5% BSA in PBST. Samples were added at a 1/50 dilution in PBST + 2% BSA, followed by incubation at 37°C/1 hour. Plates were washed five times with PBST then incubated for one hour at room temperature with polyclonal anti-human IgG antibody conjugated to HRP (Promega). Plates were again washed five times with PBST followed by addition of the chromogenic substrate TMB (Sigma) for 10 minutes and reaction stopped with 1N sulphuric acid. Absorbance was read at 450nm with an ELISA microplate reader (Biochrom Asys). A positive control specimen (from a known COVID-19 patient) in simplicate and a negative control (pre-epidemic plasma sample) in triplicate were added to every assay plate for validation and cut-off determination. Results were expressed as reaction of sample optical density value divided by the assay cutoff.

### Library Preparation and Sequencing

Total RNA from SARS-CoV-2 positive samples was converted to cDNA using the SuperScript IV First-Strand Synthesis System (Thermo Fisher Scientific, USA). Viral whole-genome amplification was performed according to the <u>Artic</u> Network protocol (https://artic.network/ncov-2019) using the SARS-CoV-2 primer scheme (V3). PCR products (pool A and B) were cleaned-up separately with AmpureXP beads (Beckman Coulter, UK), and then quantified using the Qubit dsDNA High Sensitivity assay (Life Technologies, USA). Equal amounts of each pool were combined and used to construct sequencing libraries with the TruSeq DNA Nano kit (Illumina, USA). The protocol was started at the end repair reaction, and followed as described by the manufacturer without any modifications. Library QC and quantification was performed using the High Sensitivity D1000 ScreenTape Assay on the 4200 TapeStation system (Agilent, USA). Libraries were sequenced in a MiSeq System with MiSeq Reagent Kit v3 (Illumina, USA) set to obtain 2×250 bp reads.

### Sequencing data processing and analysis

Raw read sequences in FASTQ format were first pre-processed using FastQC (v0.11.4) (https://www.bioinformatics.babraham.ac.uk/projects/fastqc/) for quality control analysis. Next, we used trimmomatic v0.39 (Bolger et al. 2014) for filtering low-quality reads, keeping those with an average quality ≥ 25. Optical duplicates in 5’ primer regions were removed using cutadapt v2.1 (Martin 2011) and clumpify v38.41 (https://sourceforge.net/projects/bbmap/), respectively. The sequences were then mapped to the Wuhan-Hu-1 reference genome (NC_045512.2) using the BWA 0.7.17 software (Li and Durbin 2009; Martin 2011). Post-processing steps were performed with samtools v1.10 (Li et al. 2009) and picard v2.17.0 packages (http://broadinstitute.github.io/picard/). We also performed *de novo* assembly using megahit programs. v.1.1.4 (Li et al. 2015) and skesa v2.4.0 (Souvorov et al. 2018).

High-frequency genetic variants were detected using the GATK v4.1.7.0 (DePristo et al. 2011) following best practices such as base recalibration using the variant database NextStrain and variant filtration (Van der Auwera et al. 2013). After variant calling, a consensus genome sequence was generated using bcftools v1.10.2 and bedtools v2.29.2 packages (Quinlan and Hall 2010; Li 2011a; Li 2011b). Low-frequency variants were called using LoFreq v 2.1.5 (Wilm et al. 2012). Next, we combined the GATK and LoFreq results and performed a pairwise variant filtration analysis using the following criteria: (i) average base quality criteria ≥ 15; (ii) allele frequency ≥ 5% and (iii) minimum coverage ≥ 50 in at least one sample of the pair. All variants were annotated using snpEff v4.5 (Cingolani et al. 2012).

### Consensus dataset collation and phylogenetic inference

We searched the GISAID database (https://www.gisaid.org) in mid-August for all complete genomes of SARS-CoV-2 collected between March and June in three Brazilian states: Rio de Janeiro (RJ), São Paulo (SP) and Minas Gerais (MG). State choice included RJ and MG due to our sampling localities and SP due to its geographic proximity and since it has a significant role in the epidemic spread of COVID-19 in Brazil. We gathered 135 genome sequences to compose our phylogenetic dataset (GISAID accession numbers are available in Table S7) and used the MAFFT algorithm to build the multiple sequence alignment from the resulting dataset (Katoh and Standley 2013). We estimated maximum likelihood phylogeny with this alignment of 201 genomes using a general time-reversible nucleotide substitution model (Tavaré 1986) with a proportion of invariable sites (GTR+I), selected by Modelfinder (Kalyaanamoorthy et al. 2017) in IQTree v.1.6.12 (Nguyen et al. 2015). Ancestral sequence reconstruction was implemented using empirical Bayesian method in IQTree v.1.6.12. Branch support values were assessed by 1000 replicates of ultrafast bootstrap approximation (Hoang et al. 2018). We assessed virus lineages for the whole dataset using Pangolin (https://pangolin.cog-uk.io) V 2.0.7 software (Rambaut et al. 2020) and checked our sequences for recombination using the full exploratory recombination method in RDP4 (Martin et al. 2015) and by the Phi-test approach (Bruen et al. 2006) in SplitsTree (Huson and Bryant 2006). No statistically significant evidence for recombination was found. We used Gblocks to select the most conserved regions of our multiple sequence alignment with default parameters (Castresana 2000). A unique block was selected with 29476 sites, representing 98% of the original alignment (flank positions: 215 – 29690).

### SARS-CoV-2 haplotype reconstruction and phylogenetic analysis

We conducted viral haplotype reconstruction from BAM and variant files that had been previously filtered for each sample. A penalized regression approach was applied using RegressHaplo based on a graph algorithm in R (Leviyang et al. 2017). Within-sample incidence- and abundance-based diversity indices were quantified using QSUtils in R (Gregori et al. 2014). We calculated indices such as richness, normalized Shannon entropy, mutation frequency by entity, functional attribute and nucleotide diversity. Alignment, lineage classification and phylogenetic inference were performed as previously described for the consensus dataset. Best fit model selected was TN+R2. We used the MEME method implemented in Datamonkey (Murrell et al. 2012) for detection of individual sites, subjected to episodic diversifying selection. An alignment including only coding regions was prepared for selection analysis. All haplotypes containing stop codons were excluded.

### Machine Learning Classification

Using the caret R package (Kuhn 2008), we built a machine learning model based on the ADA Boost algorithm (Hastie et al. 2009) optimized for tuning of parameters to evaluate classification of patient time points T1 and T2. To compose the feature matrix, we selected the following variables: viral load; missense and total variants; nucleotide substitution signatures; non-synonymous/synonymous ratio; non-coding and synonymous variants. The dataset was separated into two sets: 80% for training and 20% for testing. To evaluate model error, we applied 10,000 repeats of 10-fold cross-validation. Confusion matrix, ROC curve, and model performance metrics were calculated using the R package MLeval. Feature importance selection was applied using the caret function varImp for assigning scores and ranking features that indicate their relative importance when making a prediction. This approach has the advantage of using a model-based approach, which can incorporate the correlation structure between the predictors into the importance calculation (Huynh-Thu et al. 2012).

### Spike protein epitope analysis

To identify potential T cell S-reactive epitopes we performed an integrative analysis based on Immunology custom tracks available at UCSC Genome Browser. We downloaded and cross-referenced five different track data hubs using experimental and computational predicted T cell epitopes from T-React Epitopes (Braun et al. 2020), CD4+ and CD8+ IEDB Predictions (Grifoni et al. 2020), Poran HLA I and HLA II (Poran et al. 2020), and CD8 Rosetta MHC (Nerli and Sgourakis 2020; Poran et al. 2020). T-React Epitopes data was used as a reference resource due to the experimental validation of the peptides identified in the study. Computational predicted scores for each study were normalized using a min-max scaling feature. In each computational study, we only selected epitopes that mapped genetic variations identified in S protein. A mean score for the reference peptides was calculated in each prediction study. We then performed a binomial distribution test to compare the distribution of real peptides and the epitopes that carried mutations in our analysis.

## Data Availability

NGS data generated in our study is publicly available in SRA-NCBI (www.ncbi.nlm.nih.gov/sra), Bioproject accession PRJNA675840. Genome sequences are also deposited in Gisaid (www.gisaid.org) and the access identifiers are listed in Table S1.

## Acknowledgments

We thank the patients to taking part in this study, the Laboratório Nacional de Computação Científica - LNCC/MCTI, Universidade Federal do Rio de Janeiro and Instituto Serrapilheira.

## Workgroup Members

### Covid19-UFRJ Workgroup

Alice Laschuk Herlinger, Aliny dos Santos Carvalho, André Felipe Andrade dos Santos, Anna Carla Pinto Castiñeiras, Átila Duque Rossi, Bianca Isabelle Barreto Teixeira, Bianca Ortiz da Silva, Bruno Clarkson, Bruno Eduardo Dematté, Camila de Almeida Velozo, Camila Nacif, Camille Victória Leal Correia de Silva Caroline Macedo Nascimento, Carolyne Lalucha Alves L. da Graça, Cassia Cristina Alves Gonçalves, Cíntia Policarpo, Ekaterini Simões Goudouri, Elaine Sobral da Costa, Elisangela Costa da Silva, Enrico Bruno Riscarolli, Érica Ramos dos Santos Nascimento, Fabio Hecht Castro Medeiros, Fábio Luís Lima Monteiro, Fernanda Leitão dos Santos, Fernando Luz de Castro, Filipe Romero Rebello Moreira, Francine Bittencourt Schiffler, Gabriela Bergiante Kraychete, Gabriele Silveira da Cunha, Gisely Novaes Borges da Cunha, Guilherme Sant’Anna de Lira, Gustavo Peixoto Duarte da Silva, Harrison James Westgarth, Helena D’Anunciação de Oliveira, Helena Keito Toma, Helena Toledo Scheid, Huang Ling Fang, Inês Corrêa Gonçalves, Ingrid Camelo da Silva, Isabela Labarba Carvalho de Almeida, Jessica Maciel de Almeida, Joissy Aprigio de Oliveira, Juliana Cazarin de Menezes, Juliana Tiemi Sato Fortuna, Karyne Ferreira Monteiro, Kissyla Harley Della Pascoa França, Laura Zalcberg Renault, Lendel Correia da Costa, Leticia Averbug Correa, Liane de Jesus Ribeiro, Lídia Theodoro Boullosa, Liliane Tavares de Faria Cavalcante, Luana dos Santos Costa, Lucas Matos Millioni, Luciana Jesus da Costa, Luiza Mendonça Higa, Marcela dos Santos Durães, Marcelo Amaral de Souza, Marcelo Calado de Paula Tôrres, Mariana Freire Campos, Mariana Quinto, Mariane Talon de Menezes, Marisa Souza Correia, Mateus Rodrigues de Queiroz, Matheus Augusto Calvano Cosentino, Mayla Gabryele Miranda de Melo, Mirela D’arc Ferreira da Costa, Pedro Henrique Costa da Paz, Raissa Mirella dos Santos Cunha da Costa, Raquel Fernandes Coelho, Richard Araujo Maia, Rodrigo de Moraes Brindeiro, Romina Carvalho Ferreira, Sérgio Machado Lisboa, Thamiris dos Santos Miranda, Victoria Cortes Bastos, Viviane Guimarães Gomes.

### LNCC-Workgroup

Luciane Prioli Ciapina, Rangel Celso Souza, Éllen dos Santos Correa, Bruno Zonovelli da Silva, Amanda Araújo Serrão Andrade, Leandro Nascimento Lemos, Guilherme Cordenonsi da Fonseca.

## List of supplementary tables

**Table S1 -** Patient and sample information.

**Table S2 -** Sequencing and genome assembly features.

**Table S3 -** Intra-host single nucleotide variant (iSNV) database.

**Table S4 -** Summarization of iSNVs across sequenced samples.

**Table S5 -** Per gene and per protein mutation ratio comparison between T1 and T2.

**Table S6 -** Cross-reference mapping of Spike protein T cells and HLA epitopes modifications.

**Table S7 -** GISAID accession numbers for the populational samples and acknowledgment table.

## List of supplementary figures

**Figure S1 -** Differential genetic variation enrichment over time.

**Figure S2 -** Mutational signature frequency comparison.

**Figure S3 -** Mutational signature correlation analysis

**Figure S4 -** Long-term infection sample classification using machine learning model.

**Figure S5 -** Quasispecies richness and functional diversity inferences.

**Figure S6 -** Distribution of haplotypes frequencies between T1 and T2.

## References

Arabi YM, Mandourah Y, Al-Hameed F, Sindi AA, Almekhlafi GA, Hussein MA, Jose J, Pinto R, Al-Omari A, Kharaba A, et al. 2018. Corticosteroid Therapy for Critically Ill Patients with Middle East Respiratory Syndrome. Am. J. Respir. Crit. Care Med. 197:757–767.

Bolger AM, Lohse M, Usadel B. 2014. Trimmomatic: a flexible trimmer for Illumina sequence data. Bioinformatics [Internet] 30:2114–2120. Available from: http://dx.doi.org/10.1093/bioinformatics/btu170

Braun J, Loyal L, Frentsch M, Wendisch D, Georg P, Kurth F, Hippenstiel S, Dingeldey M, Kruse B, Fauchere F, et al. 2020. SARS-CoV-2-reactive T cells in healthy donors and patients with COVID-19. Nature [Internet]. Available from: http://dx.doi.org/10.1038/s41586-020-2598-9

Bruen TC, Philippe H, Bryant D. 2006. A simple and robust statistical test for detecting the presence of recombination. Genetics 172:2665–2681.

Castresana J. 2000. Selection of conserved blocks from multiple alignments for their use in phylogenetic analysis. Mol. Biol. Evol. 17:540–552.

CDC. 2020a. Discontinuation of Transmission-Based Precautions and Disposition of Patients with COVID-19 in Healthcare Settings (Interim Guidance). Available from: https://www.cdc.gov/coronavirus/2019-ncov/hcp/disposition-hospitalized-patients.html

CDC. 2020b. Coronavirus Disease 2019 (COVID-19). Available from: https://www.cdc.gov/coronavirus/2019-ncov/hcp/disposition-in-home-patients.html

Chester A, Somasekaram A, Tzimina M, Jarmuz A, Gisbourne J, O’Keefe R, Scott J, Navaratnam N. 2003. The apolipoprotein B mRNA editing complex performs a multifunctional cycle and suppresses nonsense-mediated decay. EMBO J. 22:3971–3982.

Cingolani P, Platts A, Wang LL, Coon M, Nguyen T, Wang L, Land SJ, Lu X, Ruden DM. 2012. A program for annotating and predicting the effects of single nucleotide polymorphisms, SnpEff. Fly [Internet] 6:80–92. Available from: http://dx.doi.org/10.4161/fly.19695

Coffin JM. 1995. HIV population dynamics in vivo: implications for genetic variation, pathogenesis, and therapy. Science 267:483–489.

DePristo MA, Banks E, Poplin R, Garimella KV, Maguire JR, Hartl C, Philippakis AA, del Angel G, Rivas MA, Hanna M, et al. 2011. A framework for variation discovery and genotyping using next-generation DNA sequencing data. Nat. Genet. 43:491–498.

Di Giorgio S, Martignano F, Torcia MG, Mattiuz G, Conticello SG. 2020. Evidence for host-dependent RNA editing in the transcriptome of SARS-CoV-2. Sci Adv 6:eabb5813.

Domingo E, Escarmís C, Sevilla N, Moya A, Elena SF, Quer J, Novella IS, Holland JJ. 1996. Basic concepts in RNA virus evolution. FASEB J. 10:859–864.

ECDC. 2020. Discharge criteria for confirmed COVID-19 cases – When is it safe to discharge COVID-19 cases from the hospital or end home isolation? European Centre for Disease Prevention and Control [Internet]. Available from: https://www.ecdc.europa.eu/sites/default/files/documents/COVID-19-Discharge-criteria.pdf Eigen M. 1993. Viral quasispecies. Sci. Am. 269:42–49.

Gomez B. 2012. Respiratory Syncytial Virus Persistence. Virology & Mycology [Internet] 01. Available from: https://www.omicsonline.org/respiratory-syncytial-virus-persistence-2161-0517.1000102.php?aid=4856

Grasselli G, Zangrillo A, Zanella A, Antonelli M, Cabrini L, Castelli A, Cereda D, Coluccello A, Foti G, Fumagalli R, et al. 2020. Baseline Characteristics and Outcomes of 1591 Patients Infected With SARS-CoV-2 Admitted to ICUs of the Lombardy Region, Italy. JAMA [Internet] 323:1574.

Available from: http://dx.doi.org/10.1001/jama.2020.5394

Greenhalgh T, Knight M, A’Court C, Buxton M, Husain L. 2020. Management of post-acute covid-19 in primary care. BMJ 370:m3026.

Gregori J, Salicrú M, Domingo E, Sanchez A, Esteban JI, Rodríguez-Frías F, Quer J. 2014. Inference with viral quasispecies diversity indices: clonal and NGS approaches. Bioinformatics 30:1104–1111.

Grifoni A, Sidney J, Zhang Y, Scheuermann RH, Peters B, Sette A. 2020. Candidate Targets for Immune Responses to 2019-Novel Coronavirus (nCoV): Sequence Homology- and Bioinformatic-Based Predictions. SSRN:3541361.

Habtemariam S, Nabavi SF, Banach M, Berindan-Neagoe I, Sarkar K, Sil PC, Nabavi SM. 2020. Should We Try SARS-CoV-2 Helicase Inhibitors for COVID-19 Therapy? Arch. Med. Res. [Internet]. Available from: http://dx.doi.org/10.1016/j.arcmed.2020.05.024

Hastie T, Rosset S, Zhu J, Zou H. 2009. Multi-class AdaBoost. Statistics and Its Interface [Internet] 2:349–360. Available from: http://dx.doi.org/10.4310/sii.2009.v2.n3.a8

Hoang DT, Chernomor O, von Haeseler A, Minh BQ, Vinh LS. 2018. UFBoot2: Improving the Ultrafast Bootstrap Approximation. Mol. Biol. Evol. 35:518–522.

Huson DH, Bryant D. 2006. Application of phylogenetic networks in evolutionary studies. Mol. Biol. Evol. 23:254–267.

Huynh-Thu VA, Saeys Y, Wehenkel L, Geurts P. 2012. Statistical interpretation of machine learning-based feature importance scores for biomarker discovery. Bioinformatics 28:1766–1774.

Jia Z, Yan L, Ren Z, Wu L, Wang J, Guo J, Zheng L, Ming Z, Zhang L, Lou Z, et al. 2019. Delicate structural coordination of the Severe Acute Respiratory Syndrome coronavirus Nsp13 upon ATP hydrolysis. Nucleic Acids Res. 47:6538–6550.

Kalyaanamoorthy S, Minh BQ, Wong TKF, von Haeseler A, Jermiin LS. 2017. ModelFinder: fast model selection for accurate phylogenetic estimates. Nat. Methods 14:587–589.

Kannan SR, Spratt AN, Quinn TP, Heng X, Lorson CL, Sönnerborg A, Byrareddy SN, Singh K. 2020. Infectivity of SARS-CoV-2: there Is Something More than D614G? J. Neuroimmune Pharmacol. [Internet]. Available from: http://dx.doi.org/10.1007/s11481-020-09954-3

Katoh K, Standley DM. 2013. MAFFT multiple sequence alignment software version 7: improvements in performance and usability. Mol. Biol. Evol. 30:772–780.

Kuhn M. 2008. Building Predictive Models in R Using the caret Package. J. Stat. Softw. 28:1–26.

Ladds E, Rushforth A, Wieringa S, Taylor S, Rayner C, Husain L, Greenhalgh T. Persistent symptoms after Covid-19: qualitative study of 114 long Covid patients and draft quality criteria for services. Available from: http://dx.doi.org/10.1101/2020.10.13.20211854

Leviyang S, Griva I, Ita S, Johnson WE. 2017. A penalized regression approach to haplotype reconstruction of viral populations arising in early HIV/SIV infection. Bioinformatics 33:2455–2463.

Li D, Liu C-M, Luo R, Sadakane K, Lam T-W. 2015. MEGAHIT: an ultra-fast single-node solution for large and complex metagenomics assembly via succinct de Bruijn graph. Bioinformatics [Internet] 31:1674–1676. Available from: http://dx.doi.org/10.1093/bioinformatics/btv033

Li H. 2011a. A statistical framework for SNP calling, mutation discovery, association mapping and population genetical parameter estimation from sequencing data. Bioinformatics 27:2987–2993.

Li H. 2011b. Improving SNP discovery by base alignment quality. Bioinformatics 27:1157–1158.

Li H, Durbin R. 2009. Fast and accurate short read alignment with Burrows-Wheeler transform. Bioinformatics 25:1754–1760.

Li H, Handsaker B, Wysoker A, Fennell T, Ruan J, Homer N, Marth G, Abecasis G, Durbin R, 1000 Genome Project Data Processing Subgroup. 2009. The Sequence Alignment/Map format and SAMtools. Bioinformatics 25:2078–2079.

Lorenzo-Redondo R, Nam HH, Roberts SC, Simons LM, Jennings LJ, Qi C, Achenbach CJ, Hauser AR, Ison MG, Hultquist JF, et al. 2020. A Unique Clade of SARS-CoV-2 Viruses is Associated with Lower Viral Loads in Patient Upper Airways. medRxiv [Internet]. Available from: http://dx.doi.org/10.1101/2020.05.19.20107144

Martin DP, Murrell B, Golden M, Khoosal A, Muhire B. 2015. RDP4: Detection and analysis of recombination patterns in virus genomes. Virus Evol 1:vev003.

Martin M. 2011. Cutadapt removes adapter sequences from high-throughput sequencing reads. EMBnet.journal [Internet] 17:10. Available from: http://dx.doi.org/10.14806/ej.17.1.200

McWilliam Leitch EC, McLauchlan J. 2013. Determining the cellular diversity of hepatitis C virus quasispecies by single-cell viral sequencing. J. Virol. 87:12648–12655.

Murrell B, Wertheim JO, Moola S, Weighill T, Scheffler K, Kosakovsky Pond SL. 2012. Detecting individual sites subject to episodic diversifying selection. PLoS Genet. 8:e1002764.

Nerli S, Sgourakis NG. 2020. Structure-based modeling of SARS-CoV-2 peptide/HLA-A02 antigens. bioRxiv [Internet]. Available from: http://dx.doi.org/10.1101/2020.03.23.004176

Nguyen L-T, Schmidt HA, von Haeseler A, Minh BQ. 2015. IQ-TREE: a fast and effective stochastic algorithm for estimating maximum-likelihood phylogenies. Mol. Biol. Evol. 32:268–274.

Niavarani A, Currie E, Reyal Y, Anjos-Afonso F, Horswell S, Griessinger E, Sardina JL, Bonnet D. 2015. APOBEC3A Is Implicated in a Novel Class of G-to-A mRNA Editing in WT1 Transcripts. PLOS ONE [Internet] 10:e0120089. Available from: http://dx.doi.org/10.1371/journal.pone.0120089

Ni M, Chen C, Qian J, Xiao H-X, Shi W-F, Luo Y, Wang H-Y, Li Z, Wu J, Xu P-S, et al. 2016. Intra-host dynamics of Ebola virus during 2014. Nat Microbiol 1:16151.

Parry J. 2020. Covid-19: Hong Kong scientists report first confirmed case of reinfection. BMJ 370:m3340.

Pavon AG, Meier D, Samim D, Rotzinger DC, Fournier S, Marquis P, Monney P, Muller O, Schwitter J. 2020. First Documentation of Persistent SARS-Cov-2 Infection Presenting With Late Acute Severe Myocarditis. Can. J. Cardiol. 36:1326.e5–e1326.e7.

Plante JA, Liu Y, Liu J, Xia H, Johnson BA, Lokugamage KG, Zhang X, Muruato AE, Zou J, Fontes-Garfias CR, et al. 2020. Spike mutation D614G alters SARS-CoV-2 fitness. Nature [Internet]. Available from: http://dx.doi.org/10.1038/s41586-020-2895-3

Poran A, Harjanto D, Malloy M, Rooney MS, Srinivasan L, Gaynor RB. 2020. Sequence-based prediction of vaccine targets for inducing T cell responses to SARS-CoV-2 utilizing the bioinformatics predictor RECON. Available from: http://dx.doi.org/10.1101/2020.04.06.027805

Prado-Vivar B, Becerra-Wong M, Guadalupe JJ, Marquez S, Gutierrez B, Rojas-Silva P, Grunauer M, Trueba G, Barragan V, Cardenas P. 2020. COVID-19 Re-Infection by a Phylogenetically Distinct SARS-CoV-2 Variant, First Confirmed Event in South America. SSRN Journal [Internet]. Available from: https://www.ssrn.com/abstract=3686174

Quinlan AR, Hall IM. 2010. BEDTools: a flexible suite of utilities for comparing genomic features. Bioinformatics 26:841–842.

Rambaut A, Holmes EC, O’Toole Á, Hill V, McCrone JT, Ruis C, du Plessis L, Pybus OG. 2020. A dynamic nomenclature proposal for SARS-CoV-2 lineages to assist genomic epidemiology. Nat Microbiol 5:1403–1407.

Richardson S, Hirsch JS, Narasimhan M, Crawford JM, McGinn T, Davidson KW, Barnaby DP, Becker LB, Chelico JD, Cohen SL, et al. 2020. Presenting Characteristics, Comorbidities, and Outcomes Among 5700 Patients Hospitalized With COVID-19 in the New York City Area. JAMA [Internet] 323:2052. Available from: http://dx.doi.org/10.1001/jama.2020.6775

Rivett L, Sridhar S, Sparkes D, Routledge M, Jones NK, Forrest S, Young J, Pereira-Dias J, Hamilton WL, Ferris M, et al. 2020. Screening of healthcare workers for SARS-CoV-2 highlights the role of asymptomatic carriage in COVID-19 transmission. Elife [Internet] 9. Available from: http://dx.doi.org/10.7554/eLife.58728

Sikkema RS, Pas SD, Nieuwenhuijse DF,O’Toole Á, Verweij J, van der Linden A, Chestakova I, Schapendonk C, Pronk M, Lexmond P, et al. 2020. COVID-19 in health-care workers in three hospitals in the south of the Netherlands: a cross-sectional study. Lancet Infect. Dis. 20:1273–1280.

Smith EC, Blanc H, Vignuzzi M, Denison MR. 2013. Coronaviruses Lacking Exoribonuclease Activity Are Susceptible to Lethal Mutagenesis: Evidence for Proofreading and Potential Therapeutics. PLoS Pathogens [Internet] 9:e1003565. Available from: http://dx.doi.org/10.1371/journal.ppat.1003565

Souvorov A, Agarwala R, Lipman DJ. 2018. SKESA: strategic k-mer extension for scrupulous assemblies. Genome Biol. 19:153.

Suárez-García I, Martínez de Aramayona López MJ, Sáez Vicente A, Lobo Abascal P. 2020. SARS-CoV-2 infection among healthcare workers in a hospital in Madrid, Spain. J. Hosp. Infect. 106:357–363.

Sun J, Xiao J, Sun R, Tang X, Liang C, Lin H, Zeng L, Hu J, Yuan R, Zhou P, et al. 2020. Prolonged Persistence of SARS-CoV-2 RNA in Body Fluids. Emerg. Infect. Dis. 26:1834–1838.

Tavaré S. 1986. Some probabilistic and statistical problems in the analysis of DNA sequences. Lectures on Mathematics in the Life Sciences 17:57–86.

To KK-W, Hung IF-N, Ip JD, Chu AW-H, Chan W-M, Tam AR, Fong CH-Y, Yuan S, Tsoi H-W, Ng AC-K, et al. 2020. Coronavirus Disease 2019 (COVID-19) Re-infection by a Phylogenetically Distinct Severe Acute Respiratory Syndrome Coronavirus 2 Strain Confirmed by Whole Genome Sequencing. Clinical Infectious Diseases [Internet]. Available from: http://dx.doi.org/10.1093/cid/ciaa1275

Van der Auwera GA, Carneiro MO, Hartl C, Poplin R, Del Angel G, Levy-Moonshine A, Jordan T, Shakir K, Roazen D, Thibault J, et al. 2013. From FastQ data to high confidence variant calls: the Genome Analysis Toolkit best practices pipeline. Curr. Protoc. Bioinformatics 43:11.10.1-11.10.33.

Vignuzzi M, Stone JK, Arnold JJ, Cameron CE, Andino R. 2006. Quasispecies diversity determines pathogenesis through cooperative interactions in a viral population. Nature 439:344–348.

Waggoner JJ, Stittleburg V, Pond R, Saklawi Y, Sahoo MK, Babiker A, Hussaini L, Kraft CS, Pinsky BA, Anderson EJ, et al. Triplex Real-Time RT-PCR for Severe Acute Respiratory Syndrome Coronavirus 2 - Volume 26, Number 7—July 2020 - Emerging Infectious Diseases journal - CDC. Available from: https://wwwnc.cdc.gov/eid/article/26/7/pdfs/20-1285.pdf

Wang Y, Guo Q, Yan Z, Zhou D, Zhang W, Zhou S, Li Y-P, Yuan J, Uyeki TM, Shen X, et al. 2018. Factors Associated With Prolonged Viral Shedding in Patients With Avian Influenza A(H7N9) Virus Infection. J. Infect. Dis. 217:1708–1717.

Wilm A, Aw PPK, Bertrand D, Yeo GHT, Ong SH, Wong CH, Khor CC, Petric R, Hibberd ML, Nagarajan N. 2012. LoFreq: a sequence-quality aware, ultra-sensitive variant caller for uncovering cell-population heterogeneity from high-throughput sequencing datasets. Nucleic Acids Res. 40:11189–11201.

Xu K, Chen Y, Yuan J, Yi P, Ding C, Wu W, Li Y, Ni Q, Zou R, Li X, et al. 2020. Factors Associated With Prolonged Viral RNA Shedding in Patients with Coronavirus Disease 2019 (COVID-19). Clin. Infect. Dis. 71:799–806.

Zhou F, Yu T, Du R, Fan G, Liu Y, Liu Z, Xiang J, Wang Y, Song B, Gu X, et al. 2020. Clinical course and risk factors for mortality of adult inpatients with COVID-19 in Wuhan, China: a retrospective cohort study. Lancet 395:1054–1062.

