## Supplementary Figures for "Intra-host evolution during SARS-CoV-2 persistent infection"

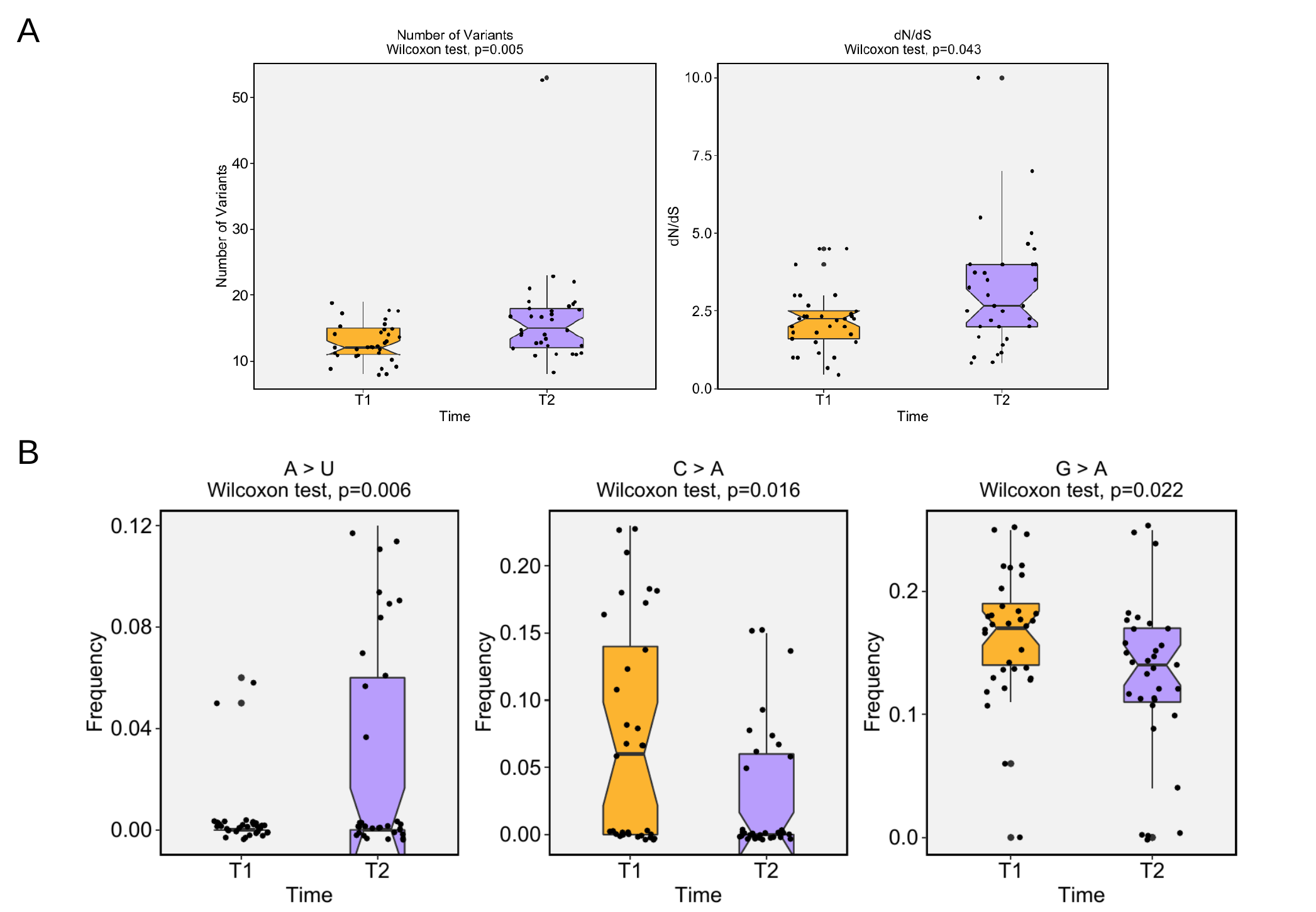

**Supplementary Figure 1. Differential genetic variation enrichment over time.** Statistical analysis of the number of variants and non-synonymous/synonymous ratio in T1 (orange) versus T2 (purple) using Wilcoxon test.

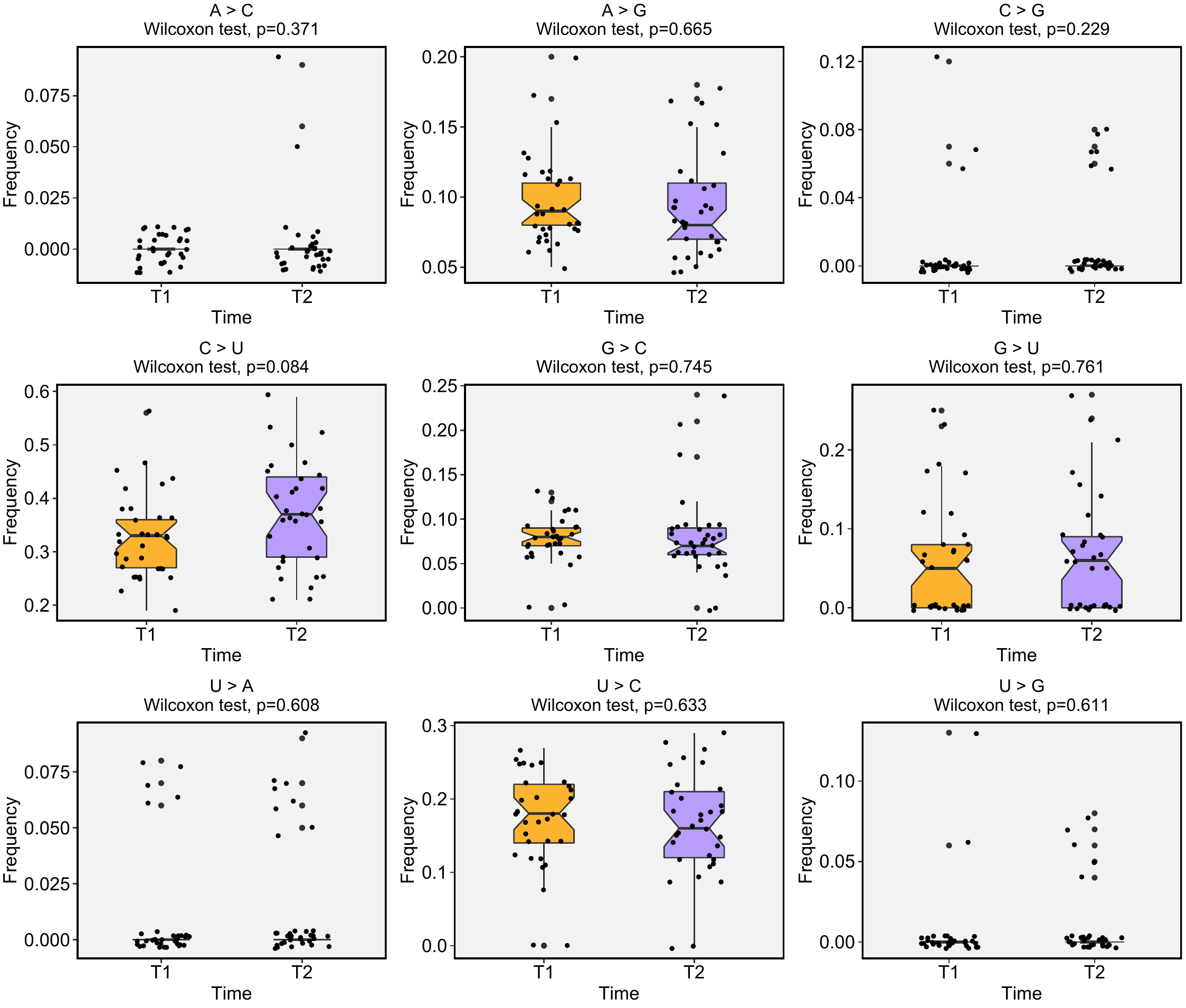

**Supplementary Figure 2. Mutational signature frequency comparison.** Statistical analysis of the transition and transversion frequencies throughout the SARS-CoV-2 genome using Wilcoxon test for each type of substitution.

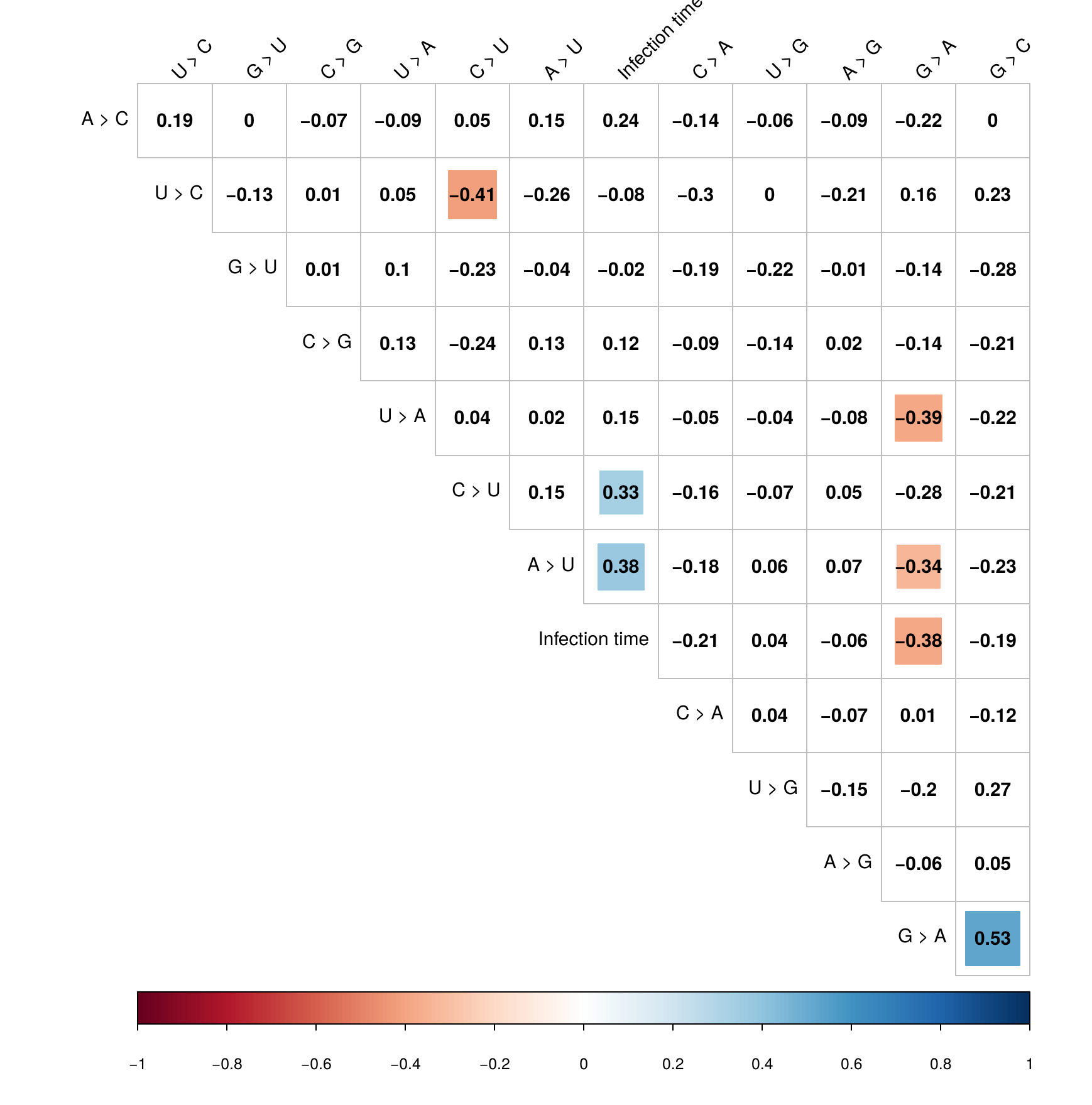

**Supplementary Figure 3. Mutational signature correlation analysis.** Pairwise Spearman’s correlation tests of the mutational signature in each patient. Color squares indicate significance level of the correlation (P < 0,05).

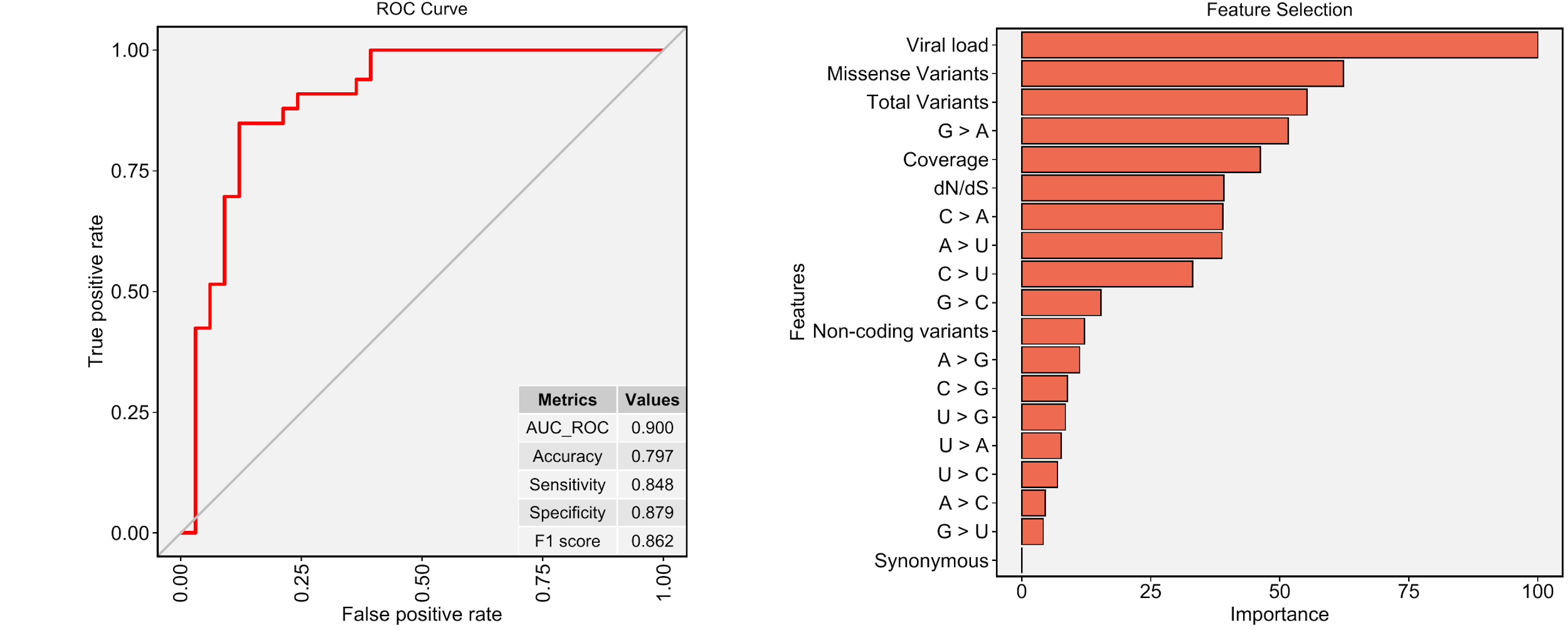

**Supplementary Figure 4. Long-term infection sample classification using machine learning model. A)** Receiver operating characteristic (ROC) curve showing a graphical representation of the relationship between sensitivity and specificity of the time point (T1 and T2) classification. The metrics table displays model performance. **B)** Overall feature importances and exhibits the most significant variables to separate the T1 and T2 classes.

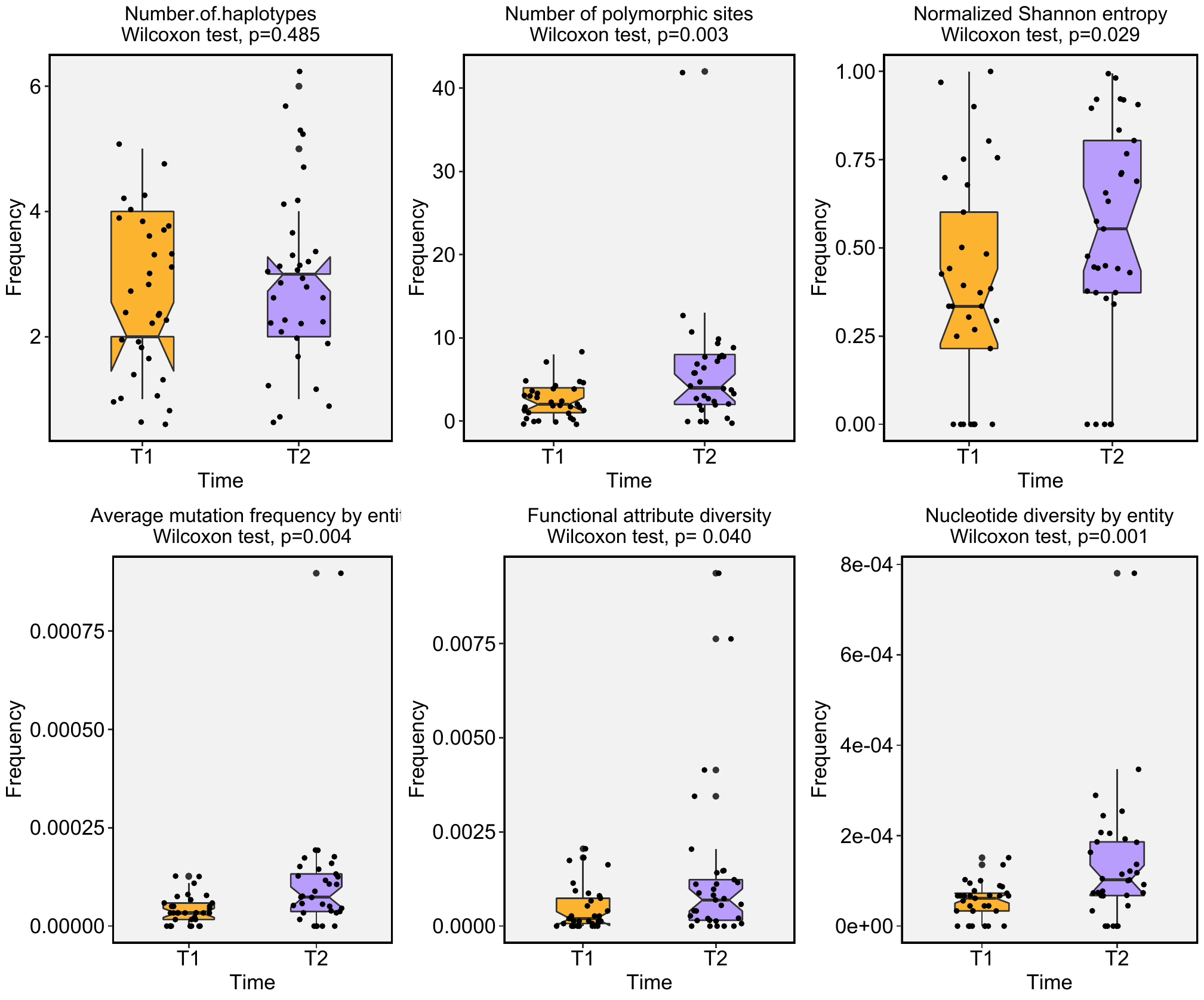

**Supplementary Figure 5. Quasispecies richness and functional diversity inferences.** Comparative analysis of the richness indices in A and B (number of haplotypes and number of polymorphic sites) and diversity (normalized Shannon entropy, average mutation frequency by entity, functional attribute diversity and nucleotide diversity by entity) between T1 and T2 samples.

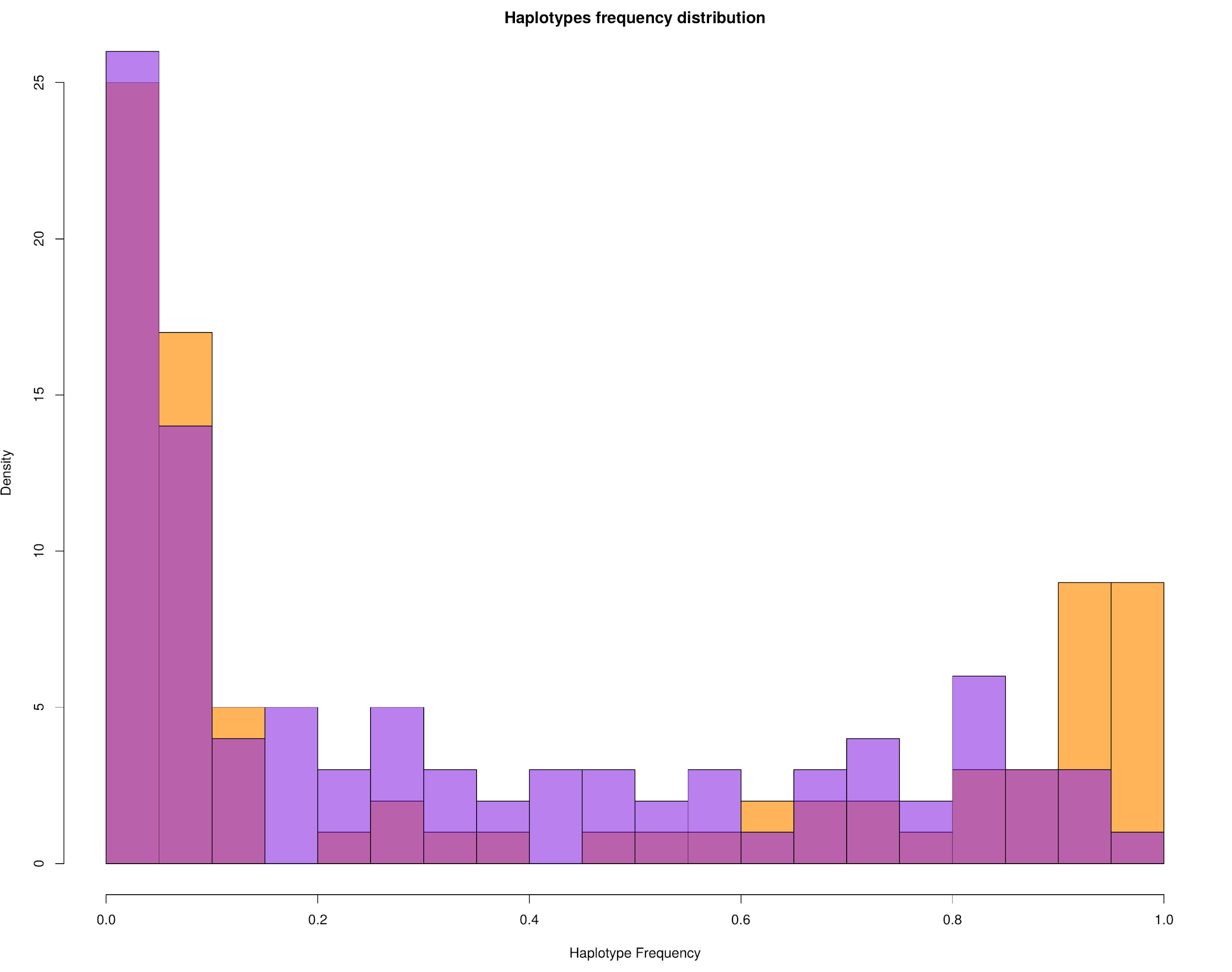

**Supplementary Figure 6. Distribution of haplotypes frequencies between T1 and T2.** Haplotypes of T1 samples are represented in orange and T2 in purple.
